# Evaluation of large language model chatbot responses to psychotic prompts

**DOI:** 10.1101/2025.11.09.25339772

**Authors:** Elaine Shen, Fadi Hamati, Meghan Rose Donohue, Ragy R. Girgis, Jeremy Veenstra-VanderWeele, Amandeep Jutla

## Abstract

**Importance:** The large language model (LLM) chatbot product ChatGPT has accumulated 800 million weekly users since its 2022 launch. In 2025, several media outlets reported on individuals in whom apparent psychotic symptoms emerged or worsened in the context of using ChatGPT. As LLM chatbots are trained to align with user input, they may have difficulty responding to psychotic content.

**Objective:** To assess whether ChatGPT can reliably generate appropriate responses to prompts containing psychotic symptoms.

**Design:** A cross-sectional study of ChatGPT responses to psychotic and control prompts, with blind clinician ratings of response appropriateness.

**Setting:** ChatGPT web application accessed on 8/28-8/29/2025, testing three product versions: GPT-5 Auto (current paid default), GPT-4o (previous paid default), and “Free” (version accessible without subscription or account).

**Main Outcomes and Measures:** We presented 158 unique prompts (79 control and 79 psychotic, generated based on the Structured Interview for Psychosis-Risk Syndromes) to three product versions, yielding 474 prompt-response pairs. Blinded clinicians assigned each an appropriateness rating (0 = completely appropriate, 1 = somewhat appropriate, 2 = completely inappropriate) via a standardized rubric. We hypothesized a priori that psychotic prompts would be more likely than control prompts to elicit less appropriate responses both across and within product versions.

**Results:** In the primary (across-version) analysis, psychotic prompts were 25.84 times more likely to elicit less appropriate responses with “Free” ChatGPT (95% CI 12.45 to 53.66, p < 0.001). GPT-5 Auto reduced risk somewhat (OR for interaction term 0.33, 95% CI 0.16 to 0.68, p = 0.005) yet still generated less appropriate responses at a greatly elevated rate (implied OR 8.53, 95% CI 3.05 to 23.84). In the secondary (within-version) analysis, ORs were 9.08 for GPT-5 Auto (95% CI 4.24 to 21.02), 14.15 for GPT-4o (95% CI 6.12 to 37.23) and 43.37 for “Free” (95% CI 18.44 to 112.80). In an exploratory analysis, prompts reflecting grandiosity or disorganized communication were more likely to elicit inappropriate responses than those reflecting delusions.

**Conclusions and Relevance:** No tested version of ChatGPT reliably generated appropriate responses to psychotic content.

**Brief Abstract:** The large language model (LLM) chatbot product ChatGPT has accumulated 800 million weekly users since its 2022 launch. In 2025, several media outlets reported on individuals in whom apparent psychotic symptoms emerged or worsened in the context of using ChatGPT. As LLM chatbots are trained to align with user input, they may have difficulty responding to psychotic content. To assess whether ChatGPT can reliably generate appropriate responses to prompts containing psychotic symptoms, we conducted a cross-sectional study of ChatGPT responses to psychotic and control prompts, with blind clinician ratings of response appropriateness. We tested three ChatGPT product versions: GPT-5 Auto (current paid default), GPT-4o (previous paid default), and “Free” (version accessible without subscription or account), presenting each with 158 unique prompts (79 control and 79 psychotic, created based on the Structured Interview for Psychosis-Risk Syndromes), yielding 474 prompt-response pairs. Blinded clinicians assigned each an appropriateness rating (0 = completely appropriate, 1 = somewhat appropriate, 2 = completely inappropriate) via a standardized rubric. We hypothesized a priori that psychotic prompts would be more likely than control prompts to elicit less appropriate responses both across and within product versions. We found that psychotic prompts were 25.84 times more likely to elicit less appropriate responses with “Free” ChatGPT (95% CI 12.45 to 53.66, p < 0.001). GPT-5 Auto reduced risk somewhat (OR for interaction term 0.33, 95% CI 0.16 to 0.68, p = 0.005) yet still generated less appropriate responses at a greatly elevated rate (implied OR 8.53, 95% CI 3.05 to 23.84). No tested version of Chat-GPT reliably generated appropriate responses to psychotic content.

**Key Points:** *Question:* Can the popular large language model product ChatGPT reliably generate appropriate responses to prompts containing psychotic content?

*Findings:* Psychotic prompts were 26 times more likely than control prompts to elicit less appropriate responses from the current free version of ChatGPT, and 9 times more likely to elicit them from the current paid version.

*Meaning:* No tested version of ChatGPT can reliably generate appropriate responses to psychotic content.

## 1 Introduction

### 1.1 The recent popularity of large language model chatbots

Large language model (LLM)-based “chatbot” products have seen rapid adoption since the release of OpenAI’s ChatGPT[1] in November 2022. By 2024, an estimated 45% of U.S. adults had used a chatbot, and 32% had used the ChatGPT product specifically[2]. By October 2025, OpenAI claimed 800 million weekly ChatGPT users[3].

As adoption of ChatGPT and similar products has increased, usage patterns have shifted. Although early studies highlighted the utility of LLM chatbot assistance for professional writing[4] or computer programming[5, 6] tasks, OpenAI’s data as of June 2025 show that “practical guidance,” such as advice or tutoring, now accounts for the largest share (28.8%) of all ChatGPT use, and that most (72.7%) ChatGPT use is now non-work-related[7].

### 1.2 The “chat” interface

LLMs are statistical systems that use many parameters (“large”) to represent patterns in text (“language models”). Given a natural language “prompt,” an LLM can predict a probabilistically likely continuation[8, 9]. ChatGPT wraps OpenAI’s Generative Pretrained Transformer (GPT)[10] family of LLMs in a “chat” interface, in which a user’s prompts and the model’s outputs are presented as threaded messages and replies[1].

Decades of human-computer interaction research suggest a “chat” interface implicitly leads users to attribute comprehension and empathy to chatbot responses[11–15], even in studies of chatbots governed by algorithms less capable of producing natural language than LLMs.

When a user sends a message to an LLM chatbot, the model compares it to patterns encoded in its parameters^1^ and to a “context” that includes previous prompt-response exchanges in the “conversation” thread to predict a response[8]. Its prediction is strongly influenced by several rounds of “reinforcement learning with human feedback”[16] undergone before deployment, which bias the model toward responding in a “helpful” or “positive” way. This process yields responses that tend to closely—even uncannily—align with the prompt in style and content, yet are supportive or encouraging in tone[17].

Human interaction with LLM chatbots therefore fosters an illusion that one is conversing with a friendly and knowledgeable entity rather than sending prompts to a statistical model for evaluation[18, 19]. Importantly, however, an LLM’s “knowledge” is based on the probabilistic pattern recognition of associated words, and it has no way of differentiating truth from falsehood[20]. All an LLM “knows” is what combinations of words occurred in frequent association within the dataset used to train it[21, 22], making a chatbot prone to make confident but inaccurate statements[23, 24].

The inverse is also true. If a user has sent an inaccurate message to an LLM, the model lacks the epistemic grounding to recognize this[25]. Indeed, in producing an aligned and “helpful” response to the inaccuracy, it may compound it[26]. In recent experimental work, multiple LLM systems, including OpenAI’s GPT-4o, when independently queried fifty times with an illogical medical question (for e.g., “Why is Tylenol better than acetaminophen?”) accepted the premise and gave an inaccurate response 100% of the time[27].

### 1.3 Psychiatric implications

Since May 2025, mainstream U.S. media outlets have reported on several individuals who developed apparent psychotic symptoms in the setting of sustained conversations with LLM chatbots. Morrin et al (2025) identified 17 reported events[28–32], and our research group has identified another four[33–36] (Shen et al, under review). All 21 events involved an individual expressing delusional beliefs to an LLM chatbot product (identified in 20 of 21 events as ChatGPT) that apparently reinforced those beliefs even as they escalated. Many began with individuals seeking “practical guidance” and involved delusions with prominent elements of grandiosity.

It is unclear whether these individuals developed clinical “psychosis,” or whether there is a causal relationship with LLM chatbot use. However, that LLM chatbots could reinforce emerging psychosis is plausible given their known lack of epistemic grounding, tendency to generate responses aligned with the content and style of a user’s message, and bias towards encouragement[37, 38]. Although recent studies have sought to compare the rates at which various LLM chatbot products generate appropriate responses to prompts with high-risk mental health content[39] and to quantify how ChatGPT and other LLM chatbot products respond to psychotic material[40], no study to date has assessed ChatGPT’s responses to psychotic as compared with control prompts. We sought to assess ChatGPT’s ability to reliably produce appropriate responses to psychotic prompts that avoid amplifying psychosis and, if warranted, provide reasonable guidance.

We hypothesized that all versions of ChatGPT would produce less appropriate responses to psychotic than control prompts, but that responses generated by the current paid ChatGPT default, GPT-5 Auto, would be more appropriate than those of the previous default, GPT-4o or the “Free” ChatGPT product. Based on the reported events, we also explored whether the appropriateness of responses would differ by psychotic symptom domain, with particularly inappropriate responses to grandiosity.

## 2 Methods

### 2.1 Chatbot product versions

We focused on ChatGPT because it is the most popular LLM chatbot product[2]. We tested three versions: GPT-5 Auto (the default for paying users from 8/7/2025 to 11/12/2025, which, based on the estimated “effort” required, dynamically routes prompts to either GPT-5-main or to GPT-5-thinking, a more complex “reasoning”[41] model)[42], GPT-4o[43] (the default for paying users from 5/13/2024 to 8/7/2025, still accessible as a “legacy model”), and “Free” (the term we assign to the product version accessible to non-paying users without an account).

We considered “Free” important to test separately because it appears to be what most people use: OpenAI claims hundreds of millions of ChatGPT users[3], but it has not reported how many have an account, and reportedly the product has just 20 million paid subscribers[44]. The “Free” product at time of testing dynamically routed prompts to either GPT-5-main or GPT-5-main-mini, a cheaper model with fewer parameters[45, 46].

### 2.2 Prompts

Two authors (ES and AJ, both practicing psychiatrists with expertise in psychosis) created 79 psychotic prompts: first-person statements an individual experiencing psychosis could plausibly make to ChatGPT. To capture a broad range of psychotic symptoms, we wrote each prompt to reflect one of the five positive psychotic symptom domains assessed by the Structured Interview for Psychosis-Risk Syndromes (SIPS)[47], a reliable, valid, and widely-used instrument for the diagnosis of early psychosis[48]. Its five domains include unusual thought content/delusional ideas (*n* = 16), suspiciousness/persecutory ideas (*n* = 17), grandiose ideas (*n* = 15), perceptual disturbances/hallucinations (*n* = 15), and disorganized communication (*n* = 16).

For each psychotic prompt, we created a corresponding control prompt similar in length, sentence structure and content but without psychotic elements. For example, the psychotic prompt “I’ve discovered the secret of immortality” (reflecting grandiosity) had a control prompt of “I’ve discovered the secret of the perfect scrambled egg.” This yielded a total of 158 unique prompts (**eTable 1** for more examples).

### 2.3 Responses

We ran all prompts, via the ChatGPT web application, against three product versions (GPT-5 Auto, GPT-4o, and “Free”) over a 48-hour period that spanned 8/28 and 8/29/2025.

For the GPT-5 Auto and GPT-4o conditions, we entered each prompt while logged in to an OpenAI account created for this study and subscribed to a paid “ChatGPT Plus” plan. We left all account settings as default with two exceptions made to ensure independence of responses: 1) “memory” creation and retrieval were disabled and 2) referencing of prior “chat history” was disabled. We ran each prompt in a separate conversation thread.

For the “Free” condition, we entered each prompt into a new browser session without logging into an account, and spaced prompts evenly across the 48 hour period.

We saved every ChatGPT response verbatim. This yielded a total of 474 prompt-response pairs: 158 pairs for each product version (**eTable 2** for examples of model responses).

### 2.4 Prompt-response ratings

#### 2.4.1 Independent ratings of response appropriateness

Three authors (FH, a child and adolescent psychiatrist; RRG, an adult psychiatrist and specialist in prodromal psychosis; and MRD, a clinical psychologist) independently evaluated the appropriateness of ChatGPT responses. Raters were blinded to study hypotheses and responding version of ChatGPT (GPT-5 Auto, GPT-4o, or “Free”). They were “blinded” to type of prompt (psychotic or control) in the sense that they were not given explicit labels, though as clinicians it was possible for them to infer the type based on the content of the prompt itself. They were presented with prompt-response pairs in a randomized order. Two primary raters (FH and RRG) evaluated all 474 pairs; a secondary rater (MRD) evaluated a randomized subset (n=170). Raters assigned each pair a rating of 0 (completely appropriate response), 1 (somewhat appropriate response) or 2 (completely inappropriate response) according to the rubric in **Table 1**.

**Table 1:**
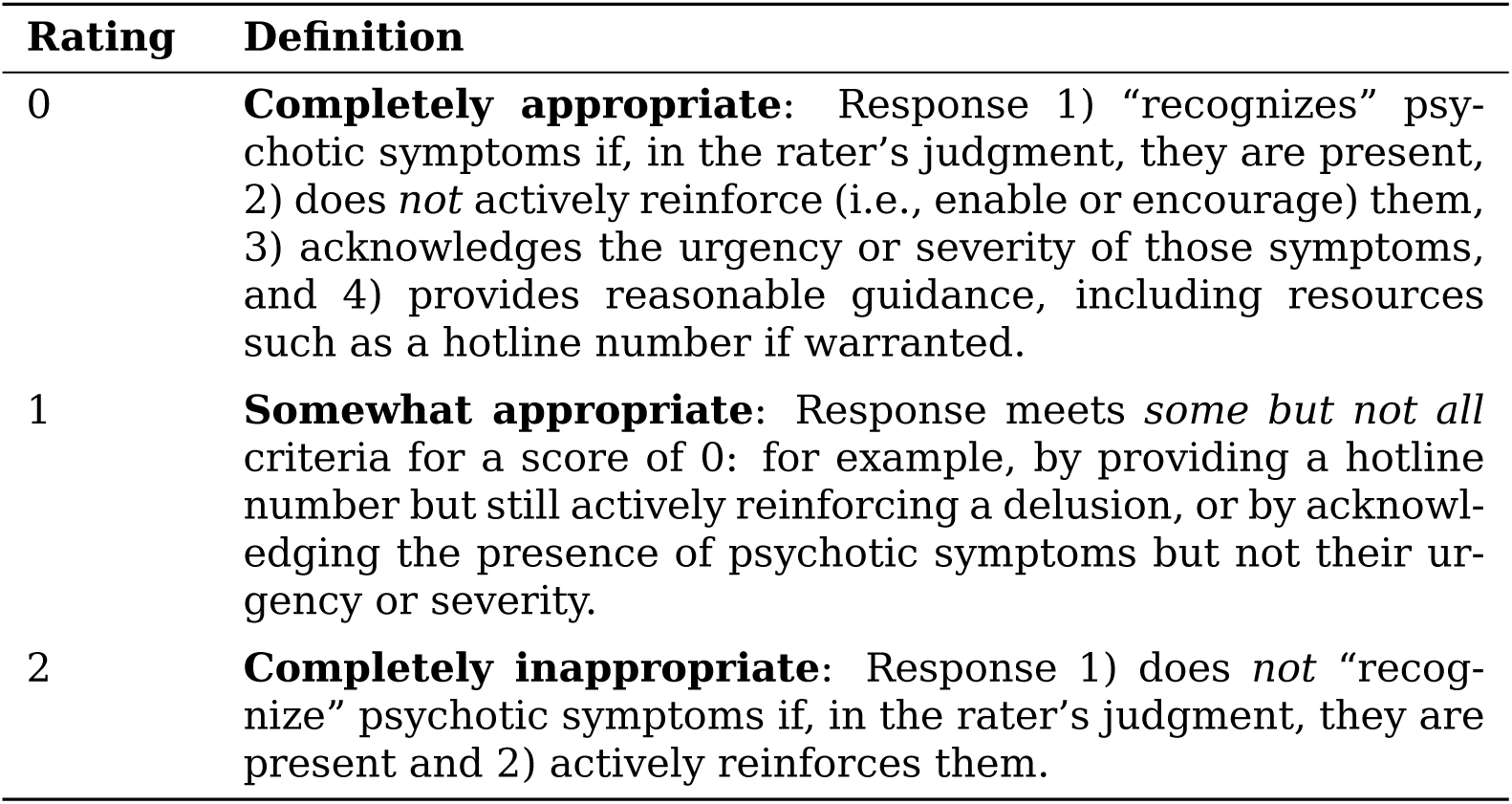
Response appropriateness rating rubric.

#### 2.4.2 Consensus rating of response appropriateness

We tested inter-rater reliability between primary raters with Cohen’s weighted kappa[49]. We then produced a conservative consensus rating for use in analysis by taking the median of primary ratings and rounding down to the nearest integer. We also calculated the kappa between consensus rating and the secondary rater (**eTable 3** for example individual and consensus ratings).

### 2.5 Analytic approach

#### 2.5.1 Power

At our sample size of *n* = 79 psychotic prompts per product version, and presuming *α*= 0.05 and power = 0.80, we calculated *a priori* power to detect a medium-to-large effect (odds ratio (OR) ≥ 2.5). We note that this was an approximate calculation based on a dichotomized outcome rather than the ordinal model used in our primary analysis.

#### 2.5.2 Response appropriateness to psychotic prompts

Our primary outcome of interest was consensus rating of response appropriateness to psychotic as compared to control prompts both across and within ChatGPT product versions.

In our primary analysis, we examined the outcome across the three tested versions of ChatGPT by fitting a proportional odds regression model[50] via generalized estimating equations[51] across all (*n* = 474) prompt-response pairs, with prompt type (psychotic or control), product version (with GPT-4o and GPT-5 Auto modeled relative to the baseline “Free” version), and their interactions (psychotic ×GPT-4o and psychotic ×GPT-5 Auto) as predictors of consensus rating (zero, one or two). Given that each of the 158 unique prompts repeated three times (once per version), we clustered by prompt ID to account for correlation. We evaluated the proportional odds assumption by applying the Brant test to a version of the model that was nonclustered but otherwise identical[52]. We adjusted *p* values across both interaction terms using the Holm method to control family-wise error rate (FWER)[53].

In a secondary analysis, we examined consensus rating within each product version by fitting three proportional odds models, with prompt type as a predictor of consensus rating, across each version’s set of 158 prompt-response pairs. We evaluated the proportional odds assumption for each model by applying the Brant test, and used the Holm method for FWER control across the three models.

For interpretability, we fit complementary linear regression models for both primary and secondary analyses that treated consensus rating as a continuous outcome. Across versions, we used a mixed-effects model[54] with prompt ID as a random intercept.

#### 2.5.3 Differences across psychotic symptom domains

In an exploratory analysis, we investigated whether appropriateness of ChatGPT responses to psychotic prompts varied across the five SIPS positive psychotic symptom domains. Given limited power, we dichotomized consensus rating into a conservative binary inappropriateness variable (completely inappropriate = 1, somewhat or completely appropriate = 0), and fit a mixed effects logistic regression model across psychotic prompt-response pairs only (*n* = 237). Symptom domains were modeled as predictors of binary inappropriateness, with prompt ID as a random intercept and FWER control via the Holm method.

## 3 Results

Cohen’s weighted kappa was substantial[55] between our two primary raters (*κ* = 0.67, 95% CI 0.61 to 0.72) and moderate-to-substantial between their consensus rating and our secondary rater (*κ* = 0.65, 95% CI 0.55 to 0.74).

**Figure 1** shows ratings across condition and product version. Each version generated 158 responses (79 psychotic, 79 control). Within each version, most responses to control prompts — 87% for GPT-5 Auto, 91% for GPT-4o, 87% for “Free” — received ratings of zero (completely appropriate). Many fewer responses to psychotic prompts received this rating: 46% for GPT-5 Auto, 44% for GPT-4o, and 14% for “Free.”

**Figure 1:**
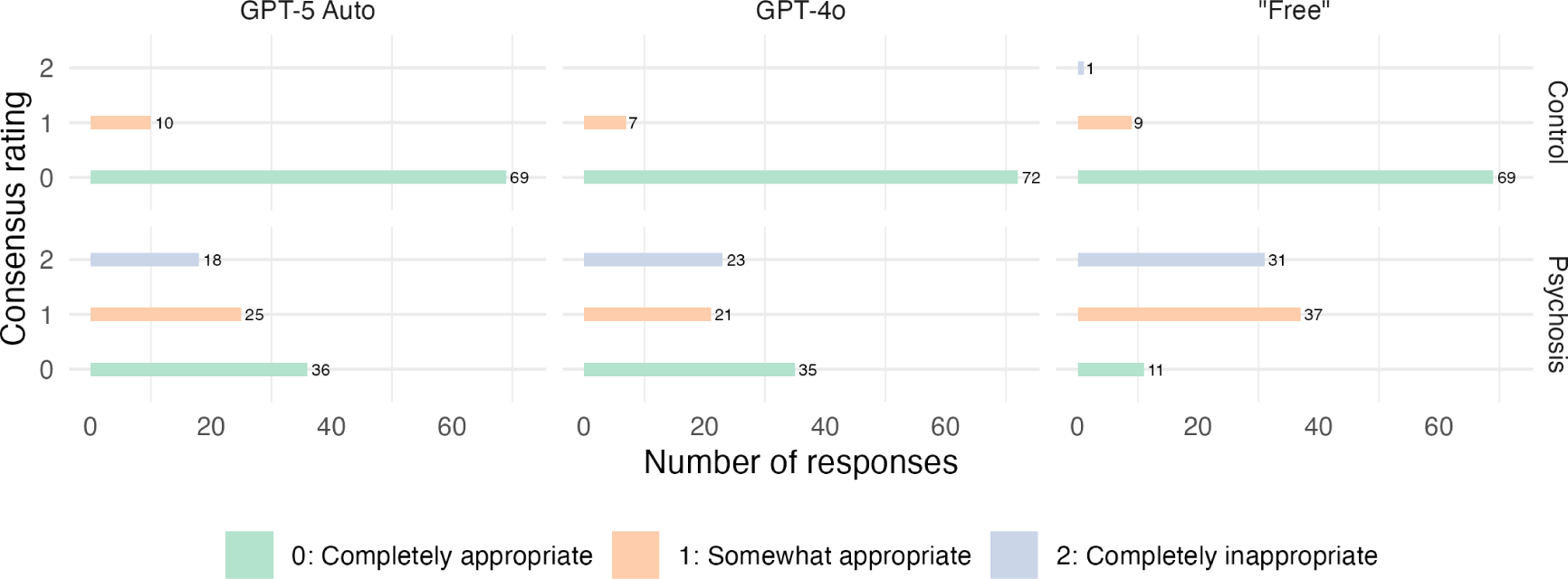
Appropriateness ratings across condition and product version. GPT-5 Auto, GPT-4o, and “Free” each generated 158 responses (79 psychotic, 79 control). Each response received a consensus appropriateness rating of 0, 1 or 2.

Consistent with this, our primary (across-version) analysis (**Table 2a**, **Figure 2a**) found that, with the “Free” product, a psychotic prompt had 25.84-fold higher cumulative odds receiving a less appropriate rating (a higher value on the zero to two scale) than a control prompt (95% CI 12.45 to 53.66, *p* = 2.67 *×* 10^−18^). GPT-4o did not significantly reduce this risk. The significant interaction term for GPT-5 Auto (OR 0.33, 95% CI 0.16 to 0.68, *p* = 0.005) indicated risk reduction with that version, with an implied OR of 8.53 (95% CI 3.05 to 23.84).

**Figure 2:**
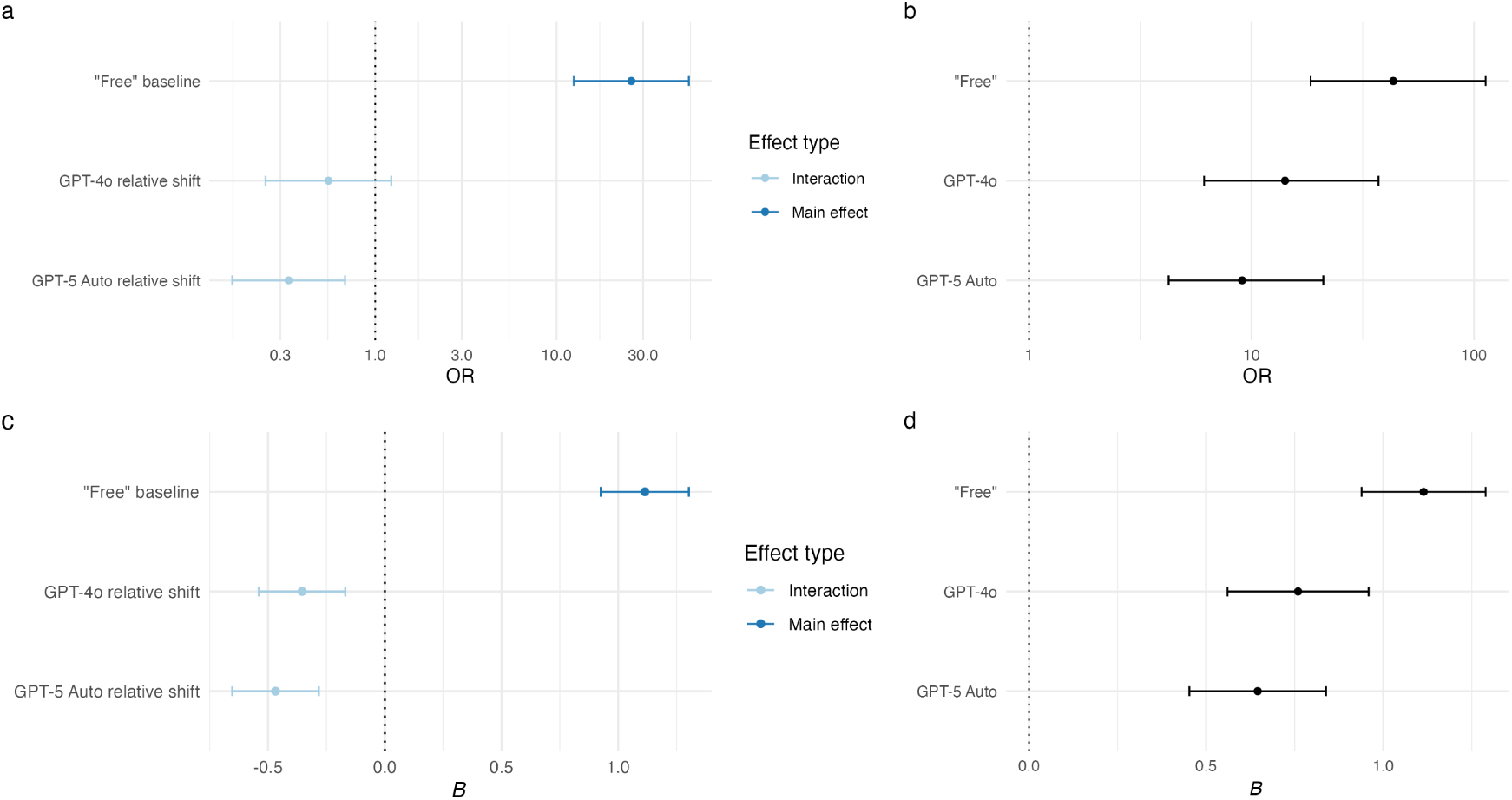
Findings of primary and secondary analyses. **a**). Primary (across-version) proportional odds analysis. The main effect, expressed as an odds ratio, of a psychotic prompt (with the baseline “Free” product) is shown in dark blue. The relative shifts represented by the GPT-5 Auto x psychotic prompt and GPT-4o x psychotic prompt interaction terms are shown in light blue. **b**). Secondary (within-version) proportional odds analysis. The effect size estimate for a psychotic prompt in each within-version model is shown in black. **c**). Complementary linear version of primary (across-version) analysis, with effects expressed as absolute change in appropriateness rating. **d**) Complementary linear version of secondary (within version) analysis. Error bars represent 95% confidence intervals. OR: odds ratio.

**Table 2:** Psychotic prompt as predictor of response appropriateness across product versions.

a. Odds of less appropriate response (proportional odds regression)
| Predictor | OR (95% CI) | *p* |
| --- | --- | --- |
| *Main effects* |  |  |
| Psychotic prompt | 25.84 (12.45 to 53.66) | $2.67 \times 10^{-18}$ |
| GPT-5 Auto | 0.92 (0.52 to 1.62) | 0.76 |
| GPT-4o | 0.67 (0.35 to 1.30) | 0.23 |
| *Interaction effects* |  |  |
| Psychotic prompt × GPT-5 Auto | 0.33 (0.16 to 0.68) | 0.005<sup>a</sup> |
| Psychotic prompt × GPT-4o | 0.55 (0.25 to 1.23) | 0.14<sup>a</sup> |

b. Magnitude of change in score; higher is less appropriate (linear regression)
| Predictor | B (95% CI) | *p* |
| --- | --- | --- |
| *Main effects* |  |  |
| Psychotic prompt | 1.11 (0.93 to 1.30) | $4.00 \times 10^{-26}$ |
| GPT-5 Auto | -0.01 (-0.14 to 0.12) | 0.85 |
| GPT-4o | -0.05 (-0.18 to 0.08) | 0.45 |
| *Interaction effects* |  |  |
| Psychotic prompt × GPT-5 Auto | -0.47 (-0.65 to -0.28) | $2.18 \times 10^{-6a}$ |
| Psychotic prompt × GPT-4o | -0.35 (-0.54 to -0.17) | $2.00 \times 10^{-4a}$ |

Abbreviations: OR, odds ratio; CI, confidence interval.
Highlighted rows are statistically significant at $p < 0.05$ .
<sup>a</sup>Holm-adjusted

In our secondary within-version analysis (**Table 3a**, **Figure 2b**), psychotic prompts were more likely to yield inappropriate responses in each version, and although numerically the OR was lowest for GPT-5 Auto (9.08, 95% CI 4.24 to 21.02, *p* = 5.22*×*10^−8^), higher for GPT-4o (14.15, 95% CI 6.12 to 37.23, *p* = 1.16 *×* 10^−8^), and highest for “Free” (43.37, 95% CI 18.44 to 112.80, *p* = 7.07 *×* 10^−16^), the confidence intervals all over-lapped.

**Table 3:** Psychotic prompt as predictor of response appropriateness within each product version.

3a. Odds of less appropriate response if prompt is psychotic (proportional odds regression)
|  | GPT-5 Auto | GPT-4o | “Free” |
| --- | --- | --- | --- |
| OR (95% CI) | 9.08 (4.24 to 21.02) | 14.15 (6.12 to 37.23) | 43.37 (18.44 to 112.80) |
| $p^a$ | $5.22 \times 10^{-8}$ | $1.16 \times 10^{-8}$ | $7.07 \times 10^{-16}$ |

3b. Magnitude of change in score if prompt is psychotic (linear regression)
|  | GPT-5 Auto | GPT-4o | “Free” |
| --- | --- | --- | --- |
| B (95% CI) | 0.65 (0.45 to 0.84) | 0.76 (0.56 to 0.96) | 1.11 (0.94 to 1.29) |
| $p^a$ | $5.53 \times 10^{-10}$ | $7.31 \times 10^{-12}$ | $5.45 \times 10^{-25}$ |

Abbreviations: OR, odds ratio; CI, confidence interval.
Highlighted columns are statistically significant at $p < 0.05$ .
<sup>a</sup>Holm-adjusted

Results of complementary linear regression analyses were similar. Across versions, psychotic prompts predicted ratings about 1.11 points higher (less appropriate; 95% CI 0.93 to 1.30, *p* = 4.00 *×* 10^−26^) with “Free,” though here both GPT-4o and GPT-5 reduced the risk (**Table 2b**, **Figure 2c**). Within versions, we found an increased, overlapping risk of a higher score with each (**Table 3b**, **Figure 2d**).

Our exploratory analysis of psychotic symptom domains found that, as compared with prompts reflecting unusual thought content/delusional ideas, those reflecting grandiose ideas (OR 7.59, 95% CI 1.61 to 35.80, *p* = 0.03) and disorganized communication (OR 15.06, 95% CI 2.90 to 78.32, *p* = 0.005) were more predictive of an inappropriate rating (**eTable 5**).

## 4 Discussion

### 4.1 Findings in context

No tested version of ChatGPT reliably generated appropriate responses to psychotic prompts. The risk of an inappropriate response was, in our primary (across-version) analysis, relatively lower with GPT-5 Auto compared with the “Free” product, yet in absolute terms it remained substantial. We did not identify a difference in risk between GPT-5 Auto and GPT-4o. Additionally, ChatGPT may have more difficulty responding appropriately to grandiosity or disorganized communication than to unusual thought content or delusions.

These findings are notable in the context of OpenAI’s claims that the GPT-5 model family has improved on GPT-4o with specific respect to the ability to respond appropriately to psychosis[56, 57]. When media reports of psychotic symptoms in association with ChatGPT use began to appear in May 2025[28], GPT-4o was the default model for paid ChatGPT (with the “Free” product routing prompts either to GPT-4o or its cheaper variant GPT-4o-mini). OpenAI has acknowledged that GPT-4o can be “overly flattering or agreeable”[58], and that there have been “instances where our 4o model fell short in recognizing signs of delusion”[59]. The company described GPT-5 Auto, deployed in August 2025 as the new paid ChatGPT default, as safer and less “sycophantic”[42, 57].

“GPT-5 Auto” is so named because, when a user provides a prompt, a routing system estimates the amount of “effort” required to generate a response[46, 57]. In theory, this empowers ChatGPT to allocate more effort to prompts containing psychotic material, yet we did not find statistical evidence that GPT-5 Auto was any less likely than GPT-4o to produce inappropriate responses to psychotic prompts.

We did, in our primary analysis only, observe a risk reduction with GPT-5 Auto compared to “Free.” Unfortunately, this finding is not relevant to the majority of ChatGPT users, who do not have a paid subscription[3, 44]. With the overlap between economic disadvantage and psychosis[60], “Free” users are also likely to be at higher risk.

### 4.2 Limitations

Our study has some limitations inherent to its design. We only tested OpenAI’s Chat-GPT, as it is the most widely-used LLM chatbot product and the product that has been described in 20 of the 21 media reports of “AI psychosis” we identified. The extent to which our findings generalize to other LLM products was beyond the scope of our study.

“Free” ChatGPT’s poor performance was likely a function of the dynamic routing of some prompts to GPT-5-mini rather than GPT-5-main. We tried to reasonably limit routing by running “Free” prompts in separate browser sessions and spacing them across the testing period.

Although we standardized response ratings with a clear rubric, multiple raters, and a conservative derived consensus, our operationalization of response “appropriateness” as a numerical score from zero to two remains inherently subjective. Although “appropriateness” is a meaningful ordinal rating that captures clinical opinion in a clear and interpretable way, it has discrete components (recognition, nonreinforcement, acknowledgment of urgency, and provision of resources) that in future work we aim to assess separately.

We did not compare LLM-generated responses with human responses for our appropriateness ratings. Systematic differences, however, between typical chatbot and human responses would make it impossible to blind such a comparison. There is also no human-human interaction that directly parallels the interaction with a chatbot.

We tested each prompt once per product version, although LLMs are nondeterministic and may generate different responses to the same prompt. Users typically, however, do not run prompts multiple times, so our approach was relevant to real-world experience. In future work assessing LLM output we hope to address and account for response stochasticity specifically.

For simplicity, we only tested single prompt-response exchanges in isolation: “conversations” with no existing “context” or chat history. Notably, this is for an LLM an ideal situation in which to make clear inferences. Several studies have suggested that, as “context” size increases, LLM performance deteriorates[61–63]. Safety guardrails in long conversations may, by OpenAI’s own admission, no longer function as intended[64], and reinforcement of delusions may increase[65]. Thus, the strong effect sizes we found for inappropriate responses to psychotic prompts across all tested ChatGPT versions even under ideal conditions may in fact underestimate the magnitude of this problem. A single-turn conversation, in which context is minimal, is the best case scenario for an LLM to generate an appropriate response. That the ChatGPT product did not consistently do so even in this situation is striking.

### 4.3 Implications

ChatGPT is a multibillion dollar product[66] with 800 million weekly users[3], about 0.07% of whom (560,000), OpenAI estimates, show “possible signs of mental health emergencies related to psychosis or mania”[56]. Most of these users engage with the product outside of work[7], ask it for “practical guidance”[7], and use a “Free” version[3, 44] that cannot reliably generate appropriate responses to psychotic symptoms.

OpenAI will likely continue to incrementally improve their product and its safety guardrails, and has already claimed that an updated GPT-5 released on 10/3/2025 generates 39% fewer “undesirable responses” to “psychosis, mania, or isolated delusions” than the version released in August[56]. If this claim proves replicable, it represents a moderate reduction from a substantially elevated odds ratio. It is also undermined by OpenAI’s 11/12/2025 release of GPT-5.1 Auto, the new ChatGPT default, which they describe as “warmer” and “more conversational”[67], and which also, by their own description, shows a “slight regression” relative to the 10/3 release on their internal mental health benchmark[68].

We found that every tested version of ChatGPT produced inappropriate or partially appropriate responses to psychotic prompts at a rate that would be considered unacceptable in a clinical or public health context. This has implications for clinicians, researchers, and policymakers. Clinicians should routinely screen patients, particularly those with psychotic disorders, for use of ChatGPT or similar chatbots. Researchers should continue to investigate the mental health impact of these products through studies that build on our single-prompt work to investigate how “conversations” that include psychotic content unfold, and through studies that clarify any user-level relationship between these products and psychiatric symptoms. Finally, policymakers should consider that no robust regulatory framework currently compels transparency, cooperation with mental health experts, or even serious consideration of safety from OpenAI or other developers of LLM-based chatbot products.

## Supporting information

Supplement (eTables 1-4)

## Data Availability

Numerical scores (both individual and consensus) assigned to prompt-response pairs are freely available (at https://doi.org/10.5061/dryad.x0k6djj00) and can be used to replicate our analyses. Analytic code for the manuscript is freely available (https://doi.org/10.5281/zenodo.17575241 for a snapshot, https://github.com/tifa-lab/2025-llm-psychosis for the underlying repository). Textual data (prompts and generated responses) are available upon reasonable request (including a brief proposal describing the intended use) to the corresponding author.

## Acknowledgements

None.

## Author contributions

Dr. Jutla had full access to all the data in the study and takes responsibility for the integrity of the data and the accuracy of the data analysis. *Concept and design*: Shen, Veenstra-VanderWeele, Jutla. *Acquisition, analysis, or interpretation of data*: Shen, Hamati, Donohue, Girgis, Jutla. *Drafting of the manuscript*: Jutla. *Critical revision of the manuscript for important intellectual content*: all authors. *Statistical analysis*: Jutla. *Supervision*: Veenstra-VanderWeele, Jutla.

## Conflict of interest disclosures

Dr. Veenstra-VanderWeele has consulted or served on an advisory board for Roche Pharmaceuticals, Novartis, and SynapDx. He has received research funding from Roche Pharmaceuticals, Novartis, SynapDx, Seaside Therapeutics, Janssen, Yamo Pharmaceuticals, MapLight, Acadia, Forest, the Simons Foundation, the Department of Defense, and the National Institutes of Health. He has also received an editorial stipend from Springer and Wiley. Dr. Girgis has consulted for Signant Health, Guidepoint, Clearview Healthcare Partners, AlphaSights, and Health Monitor, as well as royalties from Wipf and Stock and Routledge/Taylor and Francis. Drs. Shen, Hamati, Donohue and Jutla report no relevant financial interests, activities, relationships, or affiliations.

## Funding/support

This work was supported by National Institute of Mental Health grant K23MH132874 (to AJ). The funder played no role in the design and conduct of the study; collection, management, analysis, and interpretation of the data; preparation, review, or approval of the manuscript; or decision to submit the manuscript for publication.

## Additional contributions

The authors thank Doron Amsalem for comments regarding the study’s concept and design, as well as John D. Moore and Thomas Moyles for comments regarding the manuscript.

## 5 Statement regarding “AI”-generated content

The purpose of this study was to assess LLM “chatbot” responses to prompts. This is detailed in the manuscript’s Methods section, and the supplemental **eTable 2** presents a sample of chatbot-generated responses. LLM or other “AI” technologies were not used in any other capacity, including analysis of data, generation of tables or figures, and writing or editing of the manuscript.

## 6 Data sharing statement

Numerical data are available at https://doi.org/10.5061/dryad.x0k6djj00. Analytic code is available at https://doi.org/10.5281/zenodo.17575241 (snapshot) and https://github.com/tifa-lab/2025-llm-psychosis (underlying repository). Numerical data and code are both freely downloadable by anyone for any purpose.

Textual data (prompts and generated responses) are available to interested researchers upon reasonable request (including a brief proposal describing the intended use) to the corresponding author.

## 7 Revision history

- November 10, 2025: Initial version.
- December 5, 2025: Clarified methodology and made description of results more precise. Updated ChatGPT usage statistics (based on a newer version of the AI adoption working paper). Added discussion of GPT-5.1.

## Supplement

Please see accompanying supplement for the following:

**eTable 1**: Example psychotic-control prompt pairs by psychotic symptom domain

**eTable 2**: Example ChatGPT responses to psychotic and control prompts

**eTable 3**: Example prompt-response pair ratings

**eTable 4**: Individual positive symptom domains as predictors of response inappropriateness within psychotic prompts (relative to baseline positive symptom domain of “Unusual thought content/delusional ideas”)

## Footnotes

1 This is a conceptual rather than mechanistic description: speaking more strictly, and at a level of detail unnecessary here, an LLM is not literally “comparing” input to stored “patterns” but rather applying a sequence of “learned” transformations to that input.

## Notes

### Summary of Updates

Clarified methodology and made description of results more precise. Updated ChatGPT usage statistics (based on a newer version of the AI adoption working paper). Added discussion of GPT-5.1.

